# Characterization of the First Complete Genome Sequence of Yellow Fever Virus (YFV) isolate in Sierra Leone: Implications for Public Health

**DOI:** 10.64898/2025.12.02.25340656

**Authors:** John Demby Sandi, Taylor M. Brock-Fisher, Tiangay Mariama Patience Sallay Kallon, Marietou F Paye, Ibrahim Umaru Fofanah, Dolo Nosamiefan, Mohamed Saio Kamara, Aiah Joshua Teh, Alhaji Turay, Colby Wilkason, Ian Baudi, Christopher Tomkins-Tinch, Chelsea I’Anson, Jonathan E. Pekar, Al Ozonoff, Daniel Park, Christian Happi, Pardis C. Sabeti, Donald S. Grant

## Abstract

Yellow fever virus (YFV), a mosquito-borne flavivirus that causes severe hemorrhagic disease, is endemic in parts of South America and Africa, yet genomic data from Sierra Leone is lacking despite ongoing surveillance. Using hybrid-capture metagenomic sequencing, we generated a complete 10,611-bp YFV genome (98% coverage) from an adult male patient who reported to the Kailahun Government Hospital with fever and muscle pain. Phylogenetic analysis assigned the genome to West African Clade V via the YFV Nextstrain build. Compared with other West African isolates, the Sierra Leone genome contained 57 substitutions, three of which were non-synonymous (NS2b: N79S, NS3: V515I, and RdRp: A643V). Bayesian phylogenetics estimated the time to the most recent common ancestor with the closest sequences from Senegal and the Netherlands as January 14, 2001 (95% HPD: December 17, 1987 - April 28, 2009), suggesting long-standing, cryptic transmission. Together, these findings underscore the need for expanded genomic surveillance to monitor YFV spread, evolution, and potential impacts on vaccine performance.

## Background

Yellow fever virus (YFV) is the causative agent of yellow fever (YF), an acute mosquito-borne viral haemorrhagic fever disease endemic in several countries across South America, Central America, and Africa [1]. YFV is a member of the *Flaviviridae* family within the flavivirus genus, and carries a single-stranded, positive-sense RNA genome approximately 10,862 nucleotides in length [1]. The virus is genetically diverse, with seven phylogenetically and geographically distinct YFV genotypes reported to date [2]. Within Africa, there are five circulating clades: two in West Africa, and three in East and Central Africa [3].

Despite the availability of a safe and effective YFV vaccine [4], YFV remains a re-emerging vector-borne viral pathogen of major global concern, with multiple outbreaks reported each year in Africa and South America [4,5]. A recent global estimate by Wang and colleagues suggests an annual incidence of more than 86,000 cases, most occurring in sub-Saharan Africa [1]. Most confirmed YFV cases in Africa occur among unvaccinated individuals, highlighting the continued impact of low routine vaccination coverage in high-risk regions [6]. In Sierra Leone, YFV is among the epidemic-prone pathogens under routine surveillance, and based on serological testing, isolated YF cases have been reported across the country over the past 20 years [7,8]. However, the absence of genomic data from Sierra Leone limits our understanding of relative transmission dynamics and how locally circulating viruses relate to the YFV clades circulating elsewhere in Africa.

Through the SENTINEL study, a national pathogen-surveillance effort integrating clinical sample collection with standard and multiplex CRISPR-based detection and sequencing, we received clinical excess samples from individuals who presented to the Bo, Kailahun, and Kenema Government Hospitals, with fever from July 2024 to March 2025. We tested these samples for circulating epidemic-prone pathogens using the high-throughput Combinatorial Arrayed Reactions for Multiplexed Evaluation of Nucleic acids (CARMEN), a CRISPR/Cas13-based assay [9]. We then sequenced samples that tested positive by CARMEN for further molecular analysis. Here, we describe the molecular features of the first complete YFV genome sequence obtained in Sierra Leone.

## Results

### Sierra Leone’s YFV isolate is a member of Clade V

We detected YFV in an excess clinical sample from an adult male who presented to Kailahun Government Hospital in November 2024, with fever, joint pain, and muscle pain. The sample tested positive on the CARMEN CRISPR-based diagnostic platform and was selected for genomic analysis. Because this was a retrospective study, only limited clinical information was available, and follow-up was not possible once the patient left the collection area.

Based on a positive CARMEN result for YFV, we performed hybrid-capture metagenomic sequencing and generated a complete, 10,611bp (98% coverage) YFV genome with an average read depth of 33.6 (range: 7 - 99; Figure 1A). Using the YFV Nextstrain build, the isolate was assigned to Clade V, which circulates in West Africa. We assembled a contextual dataset of 73 high-quality (>90% complete) YFV genomes from Clades IV and V retrieved from NCBI and used these sequences to infer a maximum-likelihood phylogeny (Figure 1B).

**Figure 1.**
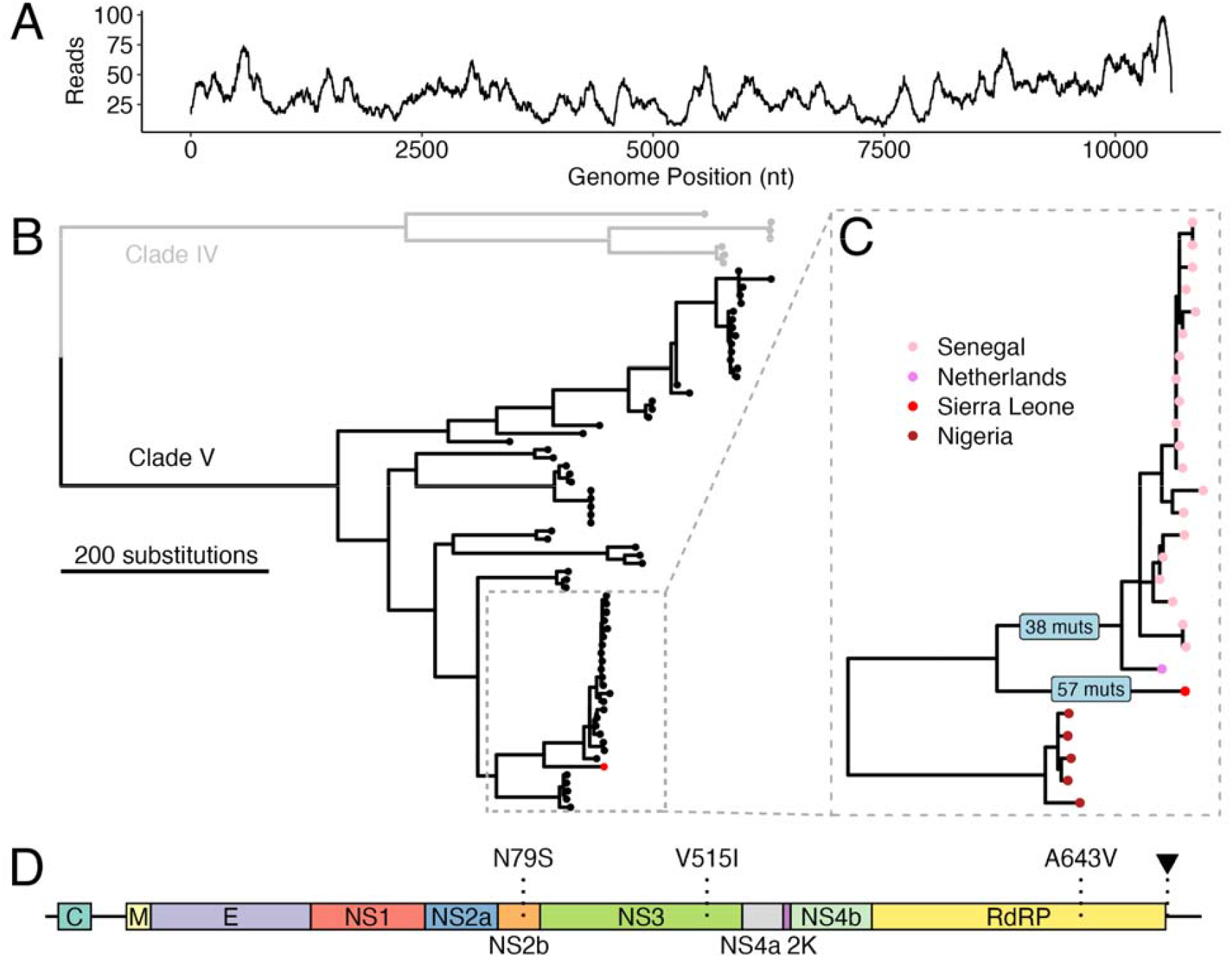
(A) Read depth profile across the 10,611-bp YFV genome generated from the Kailahun sample in Sierra Leone using hybrid-capture metagenomic sequencing. (B) Maximum likelihood tree showing the Sierra Leone YFV genome (red) in relation to a contextual dataset of 73 YFV genomes from clade IV (grey) and clade V (black). The Sierra Leone sequence clusters within Clade V. (C) Expanded view on the clade containing the Sierra Leone sequence, highlighting its position as a sister lineage to isolates from Senegal and a travel-associated case detected in the Netherlands. Numbers on branches represent the number of mutations inferred along each branch. (D) Gene map highlighting the locations of the two non-synonymous reversions (NS2b: N79S and RdRp: A643V) and one substitution (NS3: V515I) unique to the Sierra Leone YFV genome. The panel also marks the 6-nucleotide region in 3’UTR that is deleted in the Dutch and Senegalese isolates but retained in the Sierra Leone and Nigerian genomes.

Phylogenetic analysis revealed that the Sierra Leone genome formed a well supported (bootstrap support = 100) sister lineage to viruses recently detected in Senegal and a travel-associated case in the Netherlands who had reported travel in Senegal and The Gambia [10]. This combined cluster was most related to Nigeria isolates collected between 2018 and 2020 (Figure 1C). Maximum-likelihood ancestral-state reconstruction in Nextstrain [11] indicated that these sequences likely descended from a Senegalese population (confidence = 86.7%). A total of 57 mutations were inferred along the branch leading to the Sierra Leone YFV genome and 38 along the stem branch of its sister lineage; although most were synonymous. Comparisons of the Nigerian, Dutch, and Sierra Leone YFV genomes revealed two non-synonymous reversions (NS2b: N79S and RdRp: A643V) and one non-synonymous substitutions unique to the Sierra Leone genome (NS3: V515I). The Sierra Leone genome also retained the same nucleotide sequence at positions 10368-10373 of the 3’UTR as the Nigerian sample, rather than the 6-nucleotide deletion observed in the Dutch and Senegalese isolates (Figure 1D).

### YFV isolate in Sierra Leone started to diverge from the Senegalese population around mid-January 2001

Bayesian phylogenetic analysis in BEAST X [12] estimated that the Sierra Leone genome diverged from its closest related Senegalese and Dutch sequences around mid-January 2001. The median inferred time to the most recent common ancestor (tMRCA) was January 14, 2001 with a 95 percent highest posterior density (HPD) interval from December 17, 1987 to April 28, 2009 (Figure 2). The estimated evolutionary rate across the full tree was 2.49e-4 substitutions per site per year (95% HPD:1.31e-4 - 3.63e-4), consistent with previous reports for Clade V sequences [13]. These results likely support an established lineage diverging from a larger Senegalese population and persisting locally, although they cannot rule out an introduction from another undersampled region.

**Figure 2.**
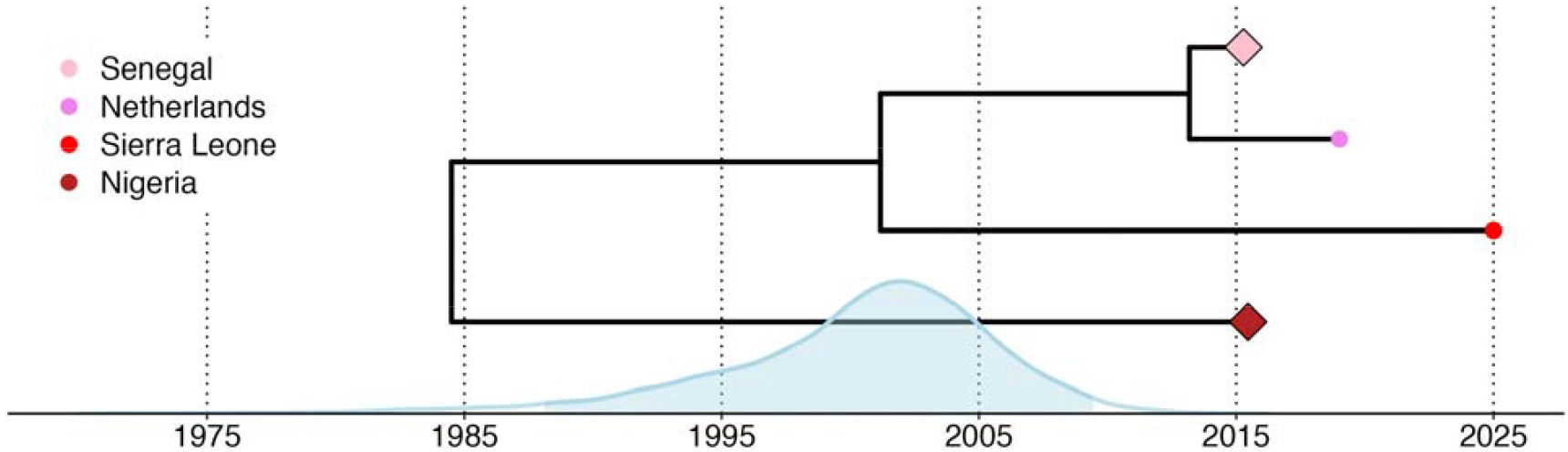
Time-scaled phylogeny inferred using BEAST X, showing the estimated divergence between the Sierra Leone genome and its closest related cluster of Senegal and Dutch genomes, as well as the Nigerian genomes. The time to the most recent common ancestor (tMRCA) is shown with the posterior height distribution, 95% HPD shaded in blue.

## Discussion

Our genomic analysis suggests that YFV has likely been present in Sierra Leone longer than previously recognized, although the limited availability of regional genomes constrains what can be concluded. The placement of the Sierra Leone genome as a sister lineage to viruses from Senegal, together with a tMRCA in January 2001, is consistent with long-standing circulation somewhere within the wider West African region. However, because YFV genomic data from Sierra Leone and neighboring countries are sparse, we cannot determine where this lineage circulated during the intervening decades or whether Sierra Leone was its primary reservoir. YFV ecology in West Africa also involves both sylvatic and zoonotic cycles, and cryptic circulation could reflect maintenance in wildlife or vectors rather than sustained human transmission. These findings underscore the need for broader genomic sampling to delineate local and regional YFV dynamics with greater confidence.

The Sierra Leone genome carries distinctive molecular features, although their functional significance remains unknown. We identified two non-synonymous reversions and one substitution not observed in closely related West African isolates, along with the retention of a 3′UTR region deleted in the Senegalese and Dutch sequences. At present, there is no evidence that these differences affect viral phenotype. Importantly, we did not detect mutations previously linked to vaccine escape and given that most confirmed YFV cases in Africa occur among unvaccinated individuals, the genomic profile of this isolate does not suggest altered vaccine sensitivity. Interpreting the impact of individual mutations is challenging without functional data, and future work will be needed to assess whether these substitutions influence viral fitness, vector interactions, or host range.

These results highlight the importance of strengthening genomic surveillance for YF in Sierra Leone and across West Africa. Routine surveillance in Sierra Leone is largely serological, which is useful for detecting exposure but insufficient for tracking viral diversity, origins, and movement. Integrating genomic sequencing into these systems would provide critical information to identify introductions, monitor lineage turnover, and evaluate potential evolutionary changes with relevance to diagnostics or vaccines. Given the re-emergence of YFV across the region, expanding genomic surveillance, paired with ecological and epidemiological data, will be essential for enabling earlier detection, improving outbreak preparedness, and informing targeted public health responses.

## Methods

### Ethics Statement

This work is part of a larger project, Surveillancing Circulating Pathogens in Sierra Leone, for which ethics approval was obtained from the Sierra Leone Ethics and Scientific Review Committee (SLESRC) (SLESRC No: 002/05/2024).

### Sampling Technique/Method

Clinical excess plasma samples collected from individuals who presented with fever at the Bo, Kailahun, and Kenema Government Hospitals were sent to the Kenema Government Hospital (KGH) viral hemorrhagic fever (VHF) research lab for a multi-viral pathogen screening as part of an ongoing diagnostic surveillance study. All samples were de-identified before transfer to the KGH VHF laboratory and were transported under cold chain conditions (2–8°C) using the standard triple packaging systems. Upon arrival at the KGH VHF laboratory, specimens were stored at −20 °C until analysis.

### Pathogen Detection

Laboratory diagnosis of pathogens of public health concern was performed using a multiplexed polymerase chain reaction (PCR) approach combined with a Combinatorial Arrayed Reactions for Multiplexed Evaluation of Nucleic acids (CARMEN) CRISPR-based detection platform, as previously described [9]. Briefly, nucleic acids were extracted from the plasma samples using the Applied Biosystems MagMAX™ Pathogen RNA/DNA Kit and subjected to a multiplex PCR amplification using in-house-designed pathogen-specific primer sets. Amplified amplicons were detected using CRISPR-Cas13, which allows for simultaneous detection of several targets in each sample on a Standard Biotools Biomark X instrument.

### Genome Sequencing

Plasma samples in which at least one pathogen was detected by CARMEN were sequenced as previously described [14]. Briefly, we used the Illumina RNA Prep with Enrichment (L) Kit and the Viral Surveillance Panel (VSP) 2.0 to generate virus-enriched libraries following the manufacturer’s protocol. The libraries were then sequenced pair-end on the Illumina MiSeq platform.

### Sierra Leone Genome Assembly

We first demultiplexed the samples using the Broad Institute demux-deplete pipeline (github.com/broadinstitute/viral-pipelines/demux_deplete), followed by genome assembly using the Broad Institute assemble-denovo-metagenomic (github.com/broadinstitute/viral-pipelines/assemble_denovo_metagenomic) in Terra.bio using default settings.

### Dataset curation and maximum likelihood phylogenetics

We used Nextstrain [11] to search NCBI for YFV genomes that were more than 90% complete (9,775 bp). The resulting genomes were downloaded and Nextclade was then used to select only Clade IV and V sequences, resulting in 73 contextual genomes (Sup. Table 1). Sequences were then aligned to the vaccine strain reference (NC.002031) using augur align. A maximum likelihood phylogeny of these samples was generated with IQ-TREE v3.0.1 [15], using a GTR substitution model and 1000 bootstrap replicates. Ancestral state reconstruction and mutation annotation were completed with augur traits and augur ancestral respectively in Nextstrain. Trees were annotated and plotted using ggtree in R [16].

### Bayesian phylogenetics

From the complete genomic dataset described above, a subset of samples from Clade V was selected to generate a time-scaled phylogeny using BEAST X [12]. We ran one chain of 100 million generations, subsampling every 10,000 steps to continuous parameter log and tree files, using a codon based substitution model (SRD06; HKY + gamma + 2) [17,18], an uncorrelated relaxed lognormal clock, and a flexible coalescent prior (a Bayesian skyride model, allowing for changes in population size through time [19]). Convergence was assessed using Tracer [20] and all ESS values were confirmed to be over 200. 10% burnin was discarded and trees were combined to an MCC tree using TreeAnnotater (https://beast.community/treeannotator) and visualized using ggtree in R.

## Data Availability

All data are available online, as detailed in Supplemental Table 1.

## Data Availability

The Sierra Leone sequence has been submitted to NCBI with accession PX571057. Code used for analysis is available from: https://github.com/broadinstitute/kgh-yellowfever.

